# HOW THE COVID-19 PANDEMIA IS SPREADING IN ITALY

**DOI:** 10.1101/2020.04.07.20056846

**Authors:** Alessandro Salvatoni

## Abstract

In this short paper we describe a simple technique aimed to forecast the course of the spreading of Covid-19 virus infection in Italy. Data released every day by the Italian “Protezione Civile” organization have been processed with reference of the the SIR classical mathematical model by *Kermack and McKendrick*. The model provides a rough estimate of the time needed to completely block virus spread. The above assumptions will be valid if Covid-19 will not be recognized as capable of establishing a chronic productive infection in significant fraction of the population.

## Background

In the last two months a progressive spreading of the SARS-CoV-2 (Covid-19) produced a world war-like scenario. Initially, most people - and probably WHO too - thought that we were facing something similar to SARS, and epidemic of 2002-2003 which caused 8,096 infected and 774 deaths in 17 countries. The problem was addressed with no great concern. Apparently, the infection started on January 2020 in China and quickly spread to South Korea, Iran and Italy. Since February 23, the Italian government, aiming to contain the infection that was causing a significant number of cases (with hospitalization rate of 10-15%) as well as cases of acute respiratory distress syndrome (ARDS) requiring ICU admissions, took drastic measures similar to those of the Chinese government. Schools were shut down, all social events were suspended, flights were progressively canceled, obligation to stay home with permissions only for stringent needs, physical and social distancing, quarantine for persons testing positive for Covid-19. Measures were gradually extended to suspension of all non-essential work activities. Smart work was stimulated. However, only people with symptoms suggestive for Covid-19 infection (fever, cough, lost of smell and taste, tiredness, dyspnea) were screened by swab. Numbers of new positives, dead and recovered people were publicly released every day by “Protezione Civile”.

One month into the above regime, an increasingly pressing question is posed by the population, i.e. how to forecast duration of the current measures. To this end, a simple mathematical model was developed that may estimate the trend of the epidemic and forecast its duration.

## Materials and Methods

For the statistical analysis of data I referred to the SIR mathematical model (*Kermack and McKendrick, 1927*). If we assume that every infected person meets another susceptible person every day, that the average duration of the disease including virus spreading is 20 days, that each person may infect an average 0f 2-3 people over 20 days, then each day an infected person can transmit the virus to 0.1-0.15 recipients. If we have 100,000 infected people, we expect that each day the infection will transmitted to 10,000-15,000 people. The survey carried out in Vo’Euganeo (a Veneto town of 3305 inhabitants) where an impressive cluster of Covid-19 infections was detected by performing PCR tests on the whole population, showed that 75% of virus-positives were asymptomatic (Preliminary unpublish data reported by A. Crisanti, 2020).

Hence, if we extend this remark to the whole nation, we may assume that the number of infected and contagious persons is three-fold that reported in Italian data of virus-positive symptomatic cases. Since recognized infectious subjects are quarantined (thus not spreading the infection), the subjects that may spread the infection are the asymptomatic ones. For example, if on day *t* the number of infected is about 90,000, about 270,000 asymptomatic contagious persons are expected. Assuming that all persons they are going meet are susceptible, about 27,000-54,000 new cases per day are expected.

According the following equation in day t:

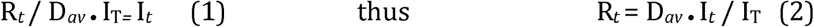

Where:

- R*t*, the R value in day *t*;
- D*av*, average duration of the contagiousness;
- I_*T*_, number of infectious subjects in day t;
- I_*t*_, number of new infected detected in day *t*

Since I_T_ subjects represent 25% of all infected (as identified by selecting symptomatic patients), the total number of Infected is I_T_ / 0.25, but only 75% of them would be contagious since not quarantined. Hence, contagious subjects would be 75% of I_T_ / 0.25. Similarly the total number of infected on day t is estimated as: I_t_ / 0.25.

Thus, taking into account also asymptomatic people (2) will be:

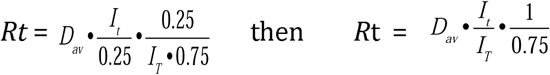

## Results

Following these considerations, the R factor was calculated on the basis of daily data reported by the Italian “Protezione Civile”: numbers of new cases, cases recovered (previous positive cases becoming virus-negative), cases that have died. The sum of the recovered and deceased cases represents the number of people that must be subtracted from the total cases for obtaining the number of actual “infectious” people each day.

The R value, upon implementation of social distancing measures, showed an exponential trend with a progressive reduction until reaching a value close to unity at the beginning of April 2020 (Figure 1). R<1 is considered the first necessary step in the control of virus spread.

**Figure 1.**
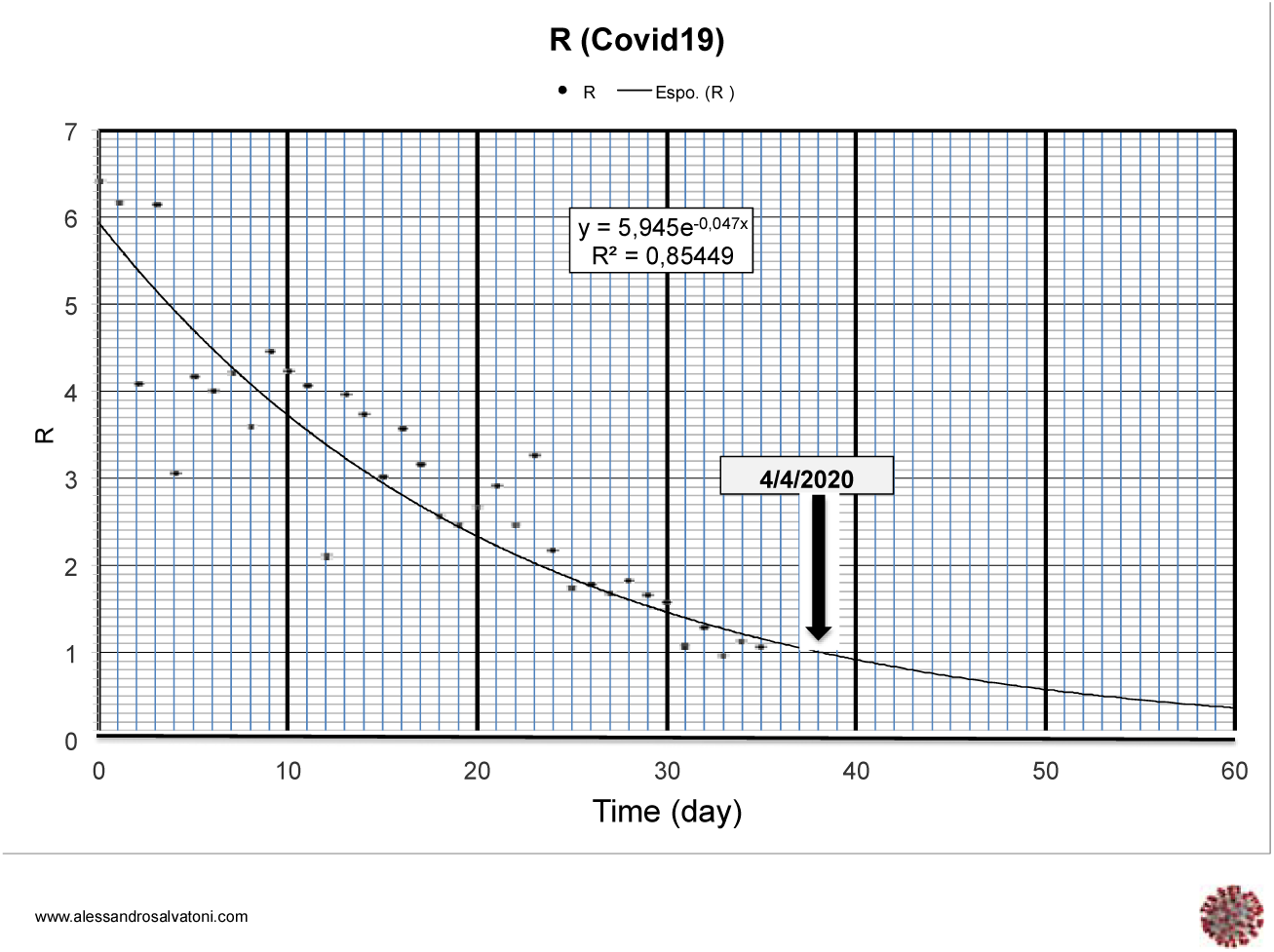
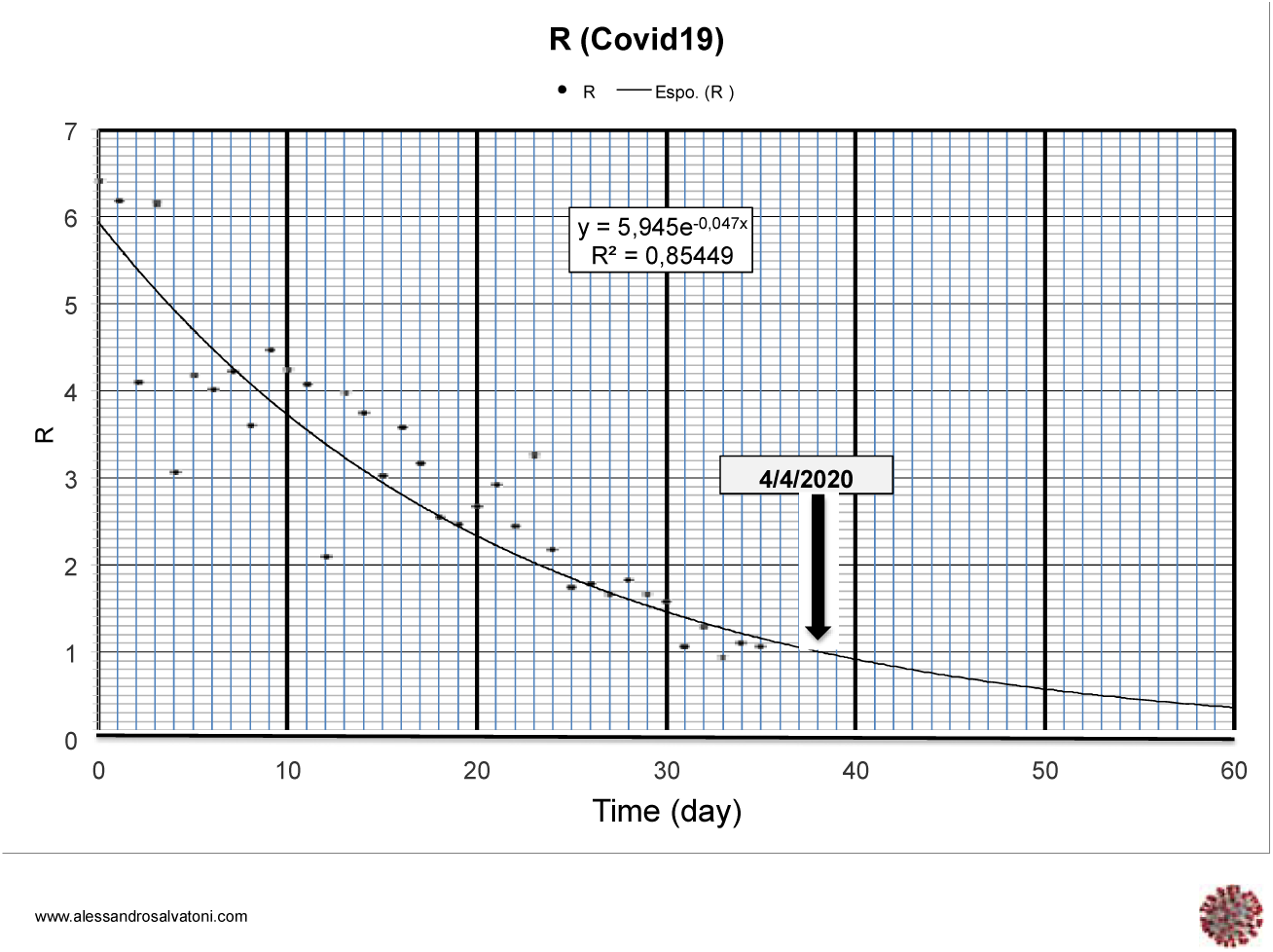
R value calculated according the formula D*I_d_/I_T_. Where D = duration of the contagiousness, I_d_ = number of new infected case for each day; I_T_=Number of total infectious subject. The exponential regression curve represents the trend of the R value.

**Figure 2.**
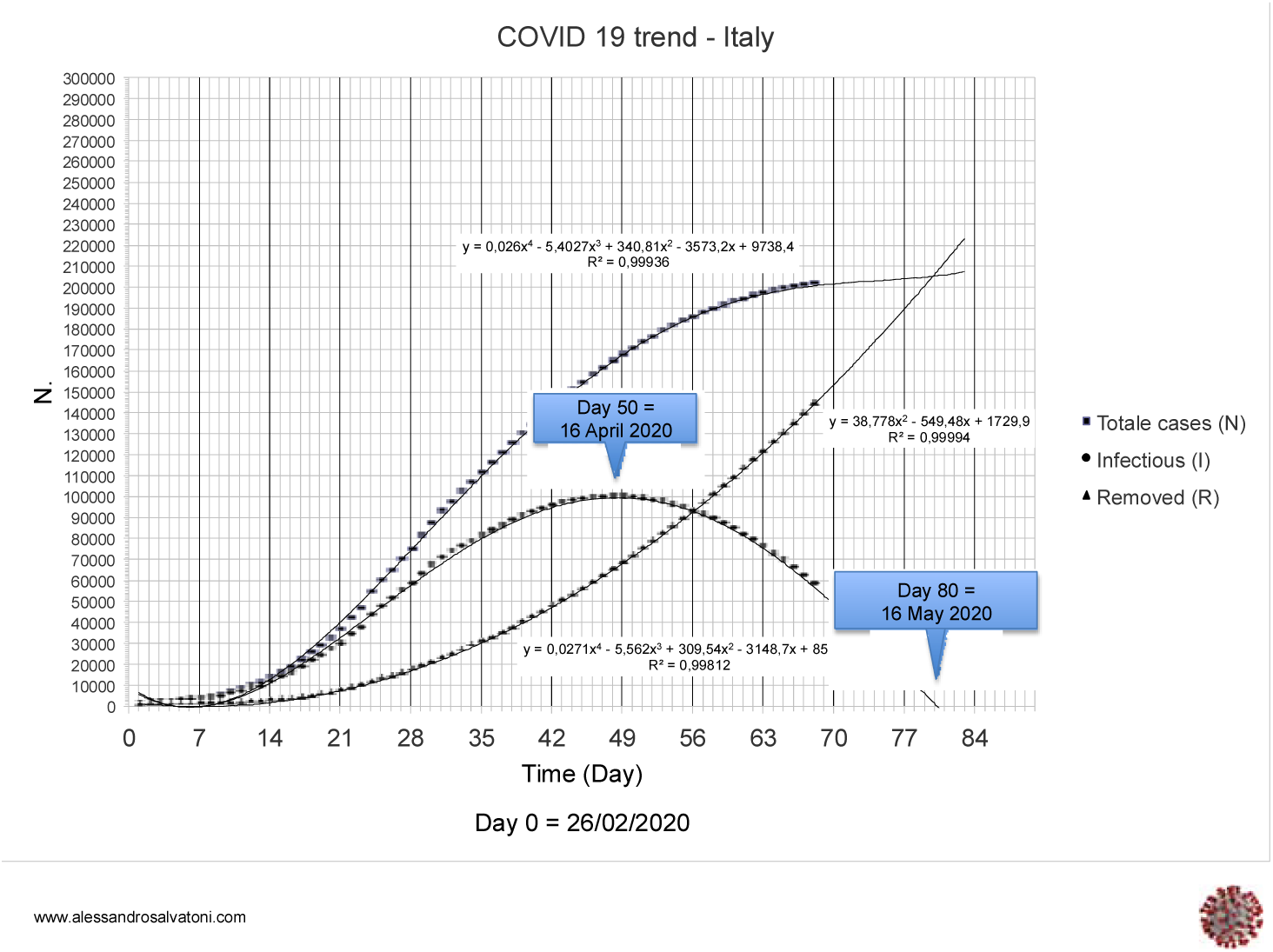
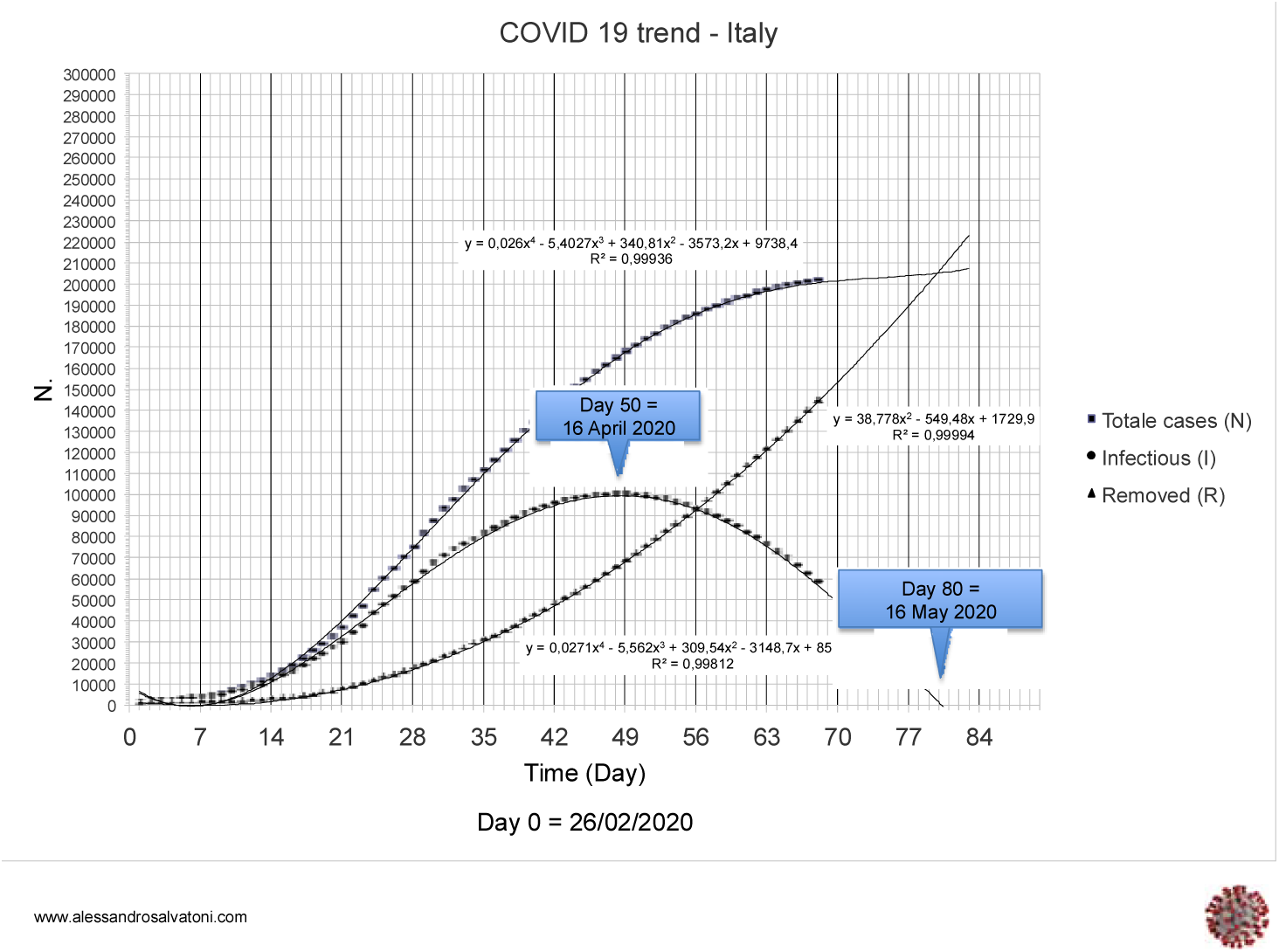
Polynomial curves, constructed on the basis of currently available data, which approximate the trend of epidemiological data relating to Covid 19 infection in Italy.

To obtain a further reduction of the contagious subjects it will take 12-14 days and to achieve the block of virus spread still 4-5 weeks.

## Discussion and conclusion

The model shows a tendency over time to overestimate the number of daily new infections. This may be due to a progressive change of non-symptomatic/symptomatic ratio, to a decrease of susceptible individual caused by social distancing measures, and/or the development of immunity by part of the population. In this contest we can estimate that the present number of asymptomatic but infectious persons roughly matches the symptomatic ones.

Detection of Covid-19 antibodies in blood of significant population samples will help answer the above questions. In this study, simplified statistical processing was used, and data were not adjusted for all possible variables influencing the course of the Covid-19 infection. However, the model provides a rough estimate of the time needed to completely block virus spread. The above assumptions will be valid if Covid-19 will not be recognized as capable of establishing a chronic productive infection in significant fraction of the population.

## Data Availability

Data are available on-line at the following address
https://lab24.ilsole24ore.com/coronavirus/

## References

Kermack WO, McKendrick AG (1927) A contribution to the mathematical theory of epidemics. Proc R Soc Lond A 115:700–721

